# Sequencing of over 100,000 individuals identifies multiple genes and rare variants associated with Crohns disease susceptibility

**DOI:** 10.1101/2021.06.15.21258641

**Authors:** Aleksejs Sazonovs, Christine R. Stevens, Guhan R. Venkataraman, Kai Yuan, Brandon Avila, Maria T. Abreu, Tariq Ahmad, Matthieu Allez, Ashwin N. Ananthakrishnan, Gil Atzmon, Aris Baras, Jeffrey C. Barrett, Nir Barzilai, Laurent Beaugerie, Ashley Beecham, Charles N. Bernstein, Alain Bitton, Bernd Bokemeyer, Andrew Chan, Daniel Chung, Isabelle Cleynen, Jacques Cosnes, David J. Cutler, Allan Daly, Oriana M. Damas, Lisa W. Datta, Noor Dawany, Marcella Devoto, Sheila Dodge, Eva Ellinghaus, Laura Fachal, Martti Farkkila, William Faubion, Manuel Ferreira, Denis Franchimont, Stacey B. Gabriel, Michel Georges, Kyle Gettler, Mamta Giri, Benjamin Glaser, Siegfried Goerg, Philippe Goyette, Daniel Graham, Eija Hämäläinen, Talin Haritunians, Graham A. Heap, Mikko Hiltunen, Marc Hoeppner, Julie E. Horowitz, Peter Irving, Vivek Iyer, Chaim Jalas, Judith Kelsen, Hamed Khalili, Barbara S. Kirschner, Kimmo Kontula, Jukka T. Koskela, Subra Kugathasan, Juozas Kupcinskas, Christopher A. Lamb, Matthias Laudes, Adam P. Levine, James Lewis, Claire Liefferinckx, Britt-Sabina Loescher, Edouard Louis, John Mansfield, Sandra May, Jacob L. McCauley, Emebet Mengesha, Myriam Mni, Paul Moayyedi, Christopher J. Moran, Rodney Newberry, Sirimon O’Charoen, David T. Okou, Bas Oldenburg, Harry Ostrer, Aarno Palotie, Joel Pekow, Inga Peter, Marieke J. Pierik, Cyriel Y. Ponsioen, Nikolas Pontikos, Natalie Prescott, Ann E. Pulver, Souad Rahmouni, Daniel L. Rice, Päivi Saavalainen, Bruce Sands, R. Balfour Sartor, Elena R. Schiff, Stefan Schreiber, L. Philip Schuum, Anthony W. Segal, Philippe Seksik, Rasha Shawky, Shehzad Z. Sheikh, Mark Silverberg, Alison Simmons, Jurgita Skeiceviciene, Harry Sokol, Matthew Solomonson, Hari Somineni, Dylan Sun, Stephan Targan, Dan Turner, Holm H. Uhlig, Andrea E. van der Meulen, Severine Vermeire, Sare Verstockt, Michiel D. Voskuil, Harland S. Winter, Justine Young, Belgium IBD Consortium, Cedars-Sinai IBD, International IBD Genetics Consortium, NIDDK IBD Genetics Consortium, NIHR IBD BioResource, Regeneron Genetics Center, SHARE Consortium, SPARC IBD Network, UK IBD Genetics Consortium, Richard H. Duerr, Andre Franke, Steven R. Brant, Judy Cho, Rinse K. Weersma, Miles Parkes, Ramnik Xavier, Manuel A. Rivas, John D. Rioux, Dermot McGovern, Hailiang Huang, Carl A. Anderson, Mark J. Daly

**Author notes:** These authors contributed equally to this work (Aleksejs Sazonovs, Christine R. Stevens, Guhan R. Venkataraman, Kai Yuan). These authors jointly supervised this work (Hailiang Huang, Carl A. Anderson, Mark J. Daly).

## Abstract

Genome-wide association studies (GWAS) have identified hundreds of loci associated with Crohns disease (CD), however, as with all complex diseases, deriving pathogenic mechanisms from these non-coding GWAS discoveries has been challenging. To complement GWAS and better define actionable biological targets, we analysed sequenced data from more than 30,000 CD patients and 80,000 population controls. We observe rare coding variants in established CD susceptibility genes as well as ten genes where coding variation directly implicates the gene in disease risk for the first time.

---

GWAS in Crohn’s Disease (CD), and inflammatory bowel disease (IBD) more generally, have successfully identified more than 200 loci contributing to risk of disease^1–4^. While most GWAS hits do not immediately implicate an obvious functional variant or gene, a subset have been directly mapped to coding variants (e.g., *NOD2, IL23R, ATG16L1, SLC39A8, FUT2, TYK2, IFIH1, SLAMF8, PLCG2*)^5^, providing more direct clues to pathogenesis. Further, targeted and genome-wide sequencing approaches have revealed additional, lower-frequency, disease-associated coding variants (e.g., *CARD9, RNF186, ADCY7, INAVA/C1orf106, SLC39A8, NOD2*)^6–9^ originally undetected by GWAS. Such coding variants, common and rare, have led to functional follow-up experiments demonstrating causal mechanisms for at least ten genes and have provided the most direct biological insights to emerge from genetic studies of IBD^10–13^.

## Results

To further advance the interpretation of GWAS loci — and to define novel CD associated genes using variation rarer than that routinely detected by GWAS — we pursued large-scale exome sequencing at multiple centers using CD case and control collections from more than 35 centers in the International IBD Genetics Consortium. The primary analysis consisted of exome sequencing of 18,828 CD cases across 35 IBD studies and 13,499 non-IBD control samples from the same studies. These samples were all sequenced at the Broad Institute and were supplemented with 22,536 population controls from approved non-IBD studies sequenced contemporaneously at the Broad Institute and accessed from dbGAP (**Supplementary Table 1**). Two different exome capture platforms were employed during the course of the study (referred to hereafter as Nextera [Illumina] and Twist [Twist Biosciences]). Details of capture and sequencing of these cohorts (and those subsequently used in follow-up) are provided in **Supplementary Material**.

Calling and quality control (QC) of data from the two exome capture platforms were conducted in parallel (**Table 1**; **Supplementary Figure 1**; **Online Methods**). Sensitivity to detect low-frequency coding variants was evaluated in each callset post-QC by comparison to passing sites in gnomAD v2.1 that had 0.0001 < non-Finnish European (NFE) minor allele frequency (MAF) < 0.1 (**Online Methods**). We observed that 84% of all exonic single nucleotide polymorphisms (SNPs) in this frequency range were detected in both CD datasets with sufficiently high quality to enter meta-analysis. Analysis of each dataset was conducted in SAIGE using a logistic mixed-model^14^, and meta-analysis was conducted with the standard inverse-variance weighted (IVW) method. Forty-three sites (**Supplementary Table 2**) failed a heterogeneity-of-effect test (IVW p_HET_ < 0.0001) and were eliminated from further analysis. We did not observe an inflation in the exome-wide distribution of test statistics (**Supplementary Figure 2**).

**Table 1.** Sample characteristics. Numbers listed are post-QC and are of samples entered into analysis. Whole-exome sequencing data derived from the UK Biobank cohort, sequenced by Regeneron using the IDT xGen Exome Research Panel v1.0 including supplemental probes, was used as population controls for the Sanger WES cases (**Online Methods**).

| Sequencing Center | Type | Exome Capture | Study stage | #CD cases | #controls |
| --- | --- | --- | --- | --- | --- |
| Broad | WES | Nextera | Discovery | 11,125 | 25,145 |
| Broad | WES | TWIST | Discovery | 6,109 | 6,064 |
| Sanger | WGS | n/a | Follow-up | 6,000 | 11,852 |
| Sanger | WES | Agilent | Follow-up | 3,731 | 33,704* |
| Regeneron | WES | Agilent | Follow-up | 4,071 | 4,223 |

Association tests were carried out for 164,149 non-synonymous variants with minor allele frequency (gnomAD NFE) between 0.0001 - 0.1, yielding a study-wide significance threshold of 3 × 10^−7^ (**Supplementary Table 3**). The most significantly-associated variants (*p* < 10^−10^) in this analysis were previously-known CD variants (or variants in linkage disequilibrium [LD] with them, **Supplementary Table 3**), indicating the QC and analysis pipeline removed highly associated false positives. Twenty-eight variants achieved study-wide significance (*p* < 3 × 10^−7^), including known variants within CD genes established in prior GWAS and sequencing efforts: *NOD2, IL23R, LRRK2, TYK2, SLC39A8, IRGM* and *CARD9*. Encouraged by this, we then nominated a list of 116 variants (including known variants) with *p* < 0.0002 for further evaluation.

Additional exome and genome sequencing was undertaken at the Sanger Institute on an independent cohort of 9,731 CD cases ascertained by the UK IBD Genetics Consortium and IBD BioResource. Genome sequencing with a target depth of 15x was performed on 6,000 CD patients. Whole-genome sequences from 11,852 individuals from the INTERVAL blood donor cohort were used as population controls. Another 3,731 CD patients were exome sequenced using the Agilent SureSelect Human All Exon V5 capture. 33,704 individuals without IBD or other related diseases from the UK Biobank were used as controls for the Sanger WES cases. These UK Biobank samples were sequenced by Regeneron using the IDT xGen Exome Research Panel v1.0 (including supplemental probes), and thus QC and subsequent analyses were restricted to the intersection of the Agilent and the IDT capture regions. Exome and genome datasets were processed in parallel with similar QC parameters (**Online Methods**).

Association analyses were performed using a logistic mixed-effects model implemented in the REGENIE software, correcting for the case-control imbalance using the Firth correction. 28 of 116 variants were associated (*p* < 4.3 × 10^−4^ (.05/116)) with CD in the meta-analysis of the two Sanger cohorts and 94 replicated the direction of effect seen in the discovery cohort (*p* = 3.34 × 10^−12^, binomial test). Summary statistics from a German dataset of 4,071 CD cases and 4,223 controls exome-sequenced at Regeneron (**Online Methods**) were also ascertained and an inverse-variance weighted meta-analysis carried out across all five cohorts (**Table 1**). 45 of the 116 variants exceeded the study-wide significance threshold, *p* < 3 × 10^−7^ (**Supplementary Table 3**).

To identify new loci not yet implicated in CD and independent exonic association signals at known loci, we accounted for the LD between these 45 variants and previously-reported IBD GWAS hits, as well as prior rare variant discoveries (**Online Methods**). We identified five coding variants in genes not previously implicated in IBD susceptibility as well as six independently associated novel exonic variants in genes previously known to harbor coding mutations underpinning CD or IBD risk, two of which are in *NOD2* (**Figure 1**; **Supplementary Table 4**).

**Figure 1.**
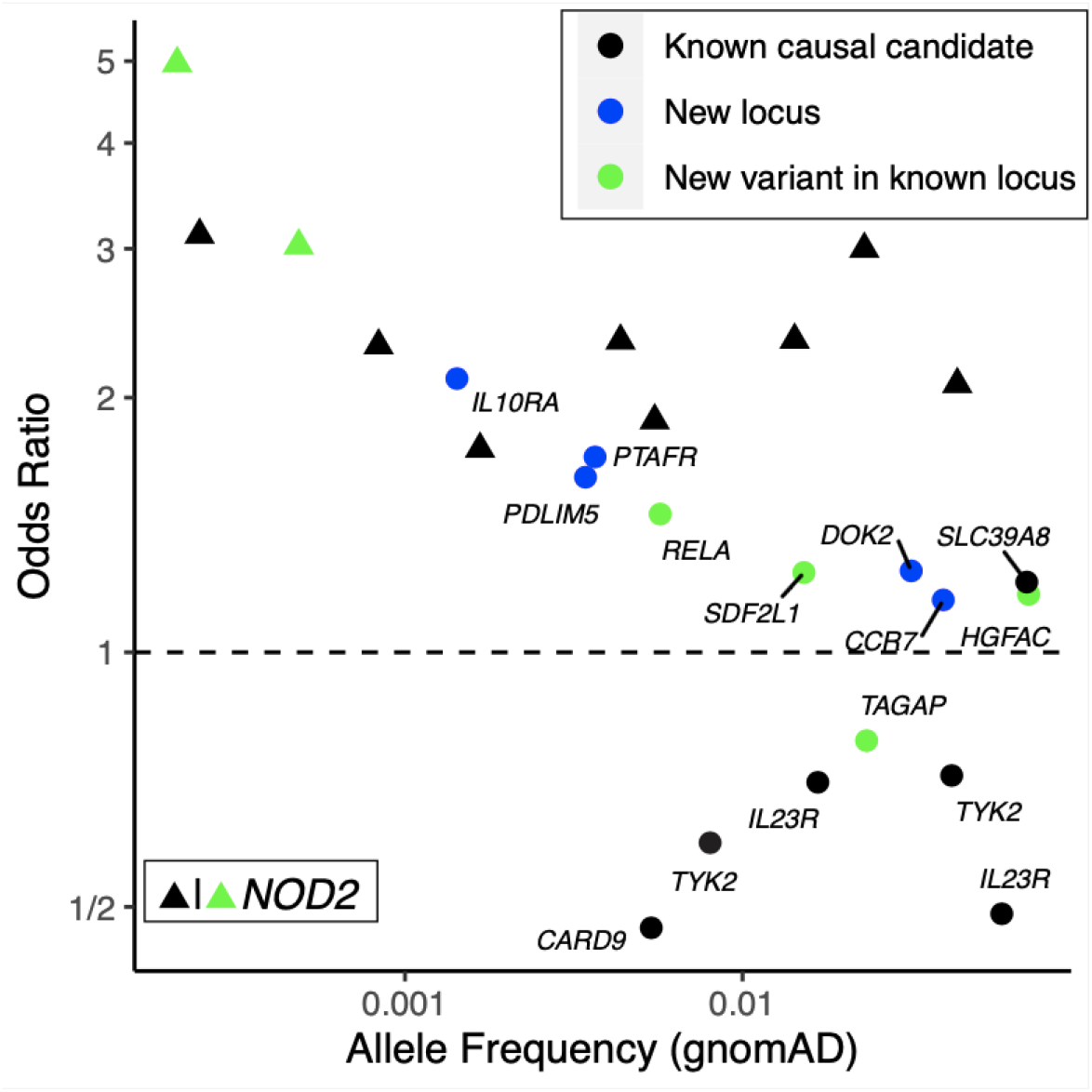
Odds ratio and minor allele frequency for exome-wide significant findings that are not tagging stronger, established non-coding association signals. *Known causal candidate:* in a credible set from a fine-mapping study^5^ with posterior inclusion probability > 5% or reported in previous studies^6,8^ (**Online Methods**). *New locus:* in a locus not yet implicated by GWAS. *New variant in known locus:* in a known GWAS locus, but represents an association independent from previously-reported IBD putative causal variants (**Online Methods**).

Most of the newly implicated CD genes (**Table 2**) play roles in biological processes already established in IBD pathogenesis, including: *DOK2*:P274L (downstream of tyrosine kinase 2: myeloid cell development and negative regulator of TLR2); *TAGAP*:E147K (Th17 differentiator and antifungal signaler); *PTAFR*:N114S (regulator of the *NLRP3* inflammasome); *CCR7*:M7V (responsible for homing of T-cells and dendritic cells, lymphocyte egress, regulatory and memory T-cell function); *IL10RA*:P295L (a regulator of innate/adaptive immune response in which recessive-acting mutations are known to cause severe neonatal enterocolitis); *RELA*:D288N (Th17 regulator with mutations reported to cause chronic mucocutaneous ulceration [CMCU]); and *HGFAC, PDLIM5* and *SDF2L1*. Further details on the newly-implicated genes are contained in **Box 1**.

**Table 2.** Variants achieving study-wide significance that implicate genes directly in general onset CD for the first time. Four of these variants (in *TAGAP*^*1,15*^, *SDF2L1*^*1*^, *RELA*^*1*^ and *HGFAC*^*2*^) are in regions highlighted in prior GWAS but represent independent associations, directly implicating these genes (**Online Methods**). Pos: genomic position in hg38; Scan *p*: *p*-value from the exome-wide discovery including subjects exome sequenced at the Broad Institute; Meta *p: p*-value from the full meta-analysis of the five cohorts shown in **Table 1**. AA Conseq.: consequence on the amino acid.

| Chr | Pos | A0 | A1 | MAF | Scan *p* | Meta *p* | OR | Gene | AA Conseq. |
| --- | --- | --- | --- | --- | --- | --- | --- | --- | --- |
| 1 | 28,150,681 | T | C | 0.0037 | 1.68E-07 | 2.96E-12 | 1.702 | *PTAFR* | N114S |
| 4 | 3,447,925 | G | A | 0.0704 | 1.37E-10 | 6.92E-15 | 1.170 | *HGFAC* | R516H |
| 4 | 94,573,345 | C | T | 0.0034 | 1.79E-05 | 2.55E-07 | 1.610 | *PDLIM5* | spl reg |
| 6 | 159,041,392 | C | T | 0.0233 | 7.60E-06 | 4.04E-10 | 0.786 | *TAGAP* | E147K |
| 8 | 21,909,729 | G | A | 0.0316 | 1.34E-04 | 2.09E-13 | 1.248 | *DOK2* | P274L |
| 11 | 65,658,293 | C | T | 0.0057 | 6.23E-05 | 2.31E-07 | 1.457 | *RELA* | D288N |
| 11 | 117,998,788 | C | T | 0.0014 | 1.13E-05 | 6.29E-09 | 2.107 | *IL10RA* | P295L |
| 17 | 40,558,934 | T | C | 0.0393 | 6.16E-06 | 4.70E-08 | 1.153 | *CCR7* | M7V/M1? |
| 22 | 21,643,991 | G | A | 0.0152 | 2.44E-05 | 2.21E-07 | 1.242 | *SDF2L1* | R161H |

The identified coding variants in *RELA, TAGAP*, and *SDF2L1* are close to, but not in LD (r^2^ < 0.05) with common non-coding variants significantly associated with IBD risk via GWAS (**Online Methods**). These very likely pinpoint the genes dysregulated by the associated common variant and provide a focus for uncovering the function of those variants, perhaps leading to allelic series of perturbations further informing on the mechanism of their contribution to CD pathogenesis. The associated missense variant in *HGFAC* is in partial LD (r2 = 0.35, in 1000 Genomes non-Finnish European populations) with a common non-coding variant (rs3752440) previously reported as associated with CD (PMID: 26192919). Unfortunately, the missense variant was not included in this previous study - precluding formal assessment of whether this explains the previously observed association signal or represents an independent variant directly implicating *HGFAC* - though we note the missense variant here has a higher odds-ratio and greater significance than the variant in the previous report. The two novel *NOD2* associations are not in LD with previously reported putative causal variants. One of them modestly reduces basal activity and has at least 2-fold reduction in peptidoglycan induced NF-κB response^16^. The other novel *NOD2* variant is a splice donor variant (**Supplementary Table 5**).

We performed two gene-based rare-variant (MAF < 0.001) burden tests in the full-exome Nextera and TWIST data sets using SAIGE-GENE^17^, one restricted to loss-of-function variants and another using all non-synonymous variants (**Supplementary Table 6**). *NOD2* unsurprisingly stood out far above the expected distribution (LoF_*p*_ = 7.7 × 10^−7^; NonSyn_*p*_ < 10^−16^). Only one other gene in either analysis exceeded the threshold expected once in the study by chance (*ATG4C*, NonSyn_*p*_ = 3.3 × 10^−6^). This potentially novel signal in *ATG4C* was driven by three distinct missense variants with individual *p* < 0.01 (N75S, R80H and C367Y) (**Supplementary Table 7**) along with two others with *p* < 0.05 (K371R, R381X). The *ATG4C* gene burden signal was examined in the Sanger data sets and replicated, with the meta-analysis reaching exome-wide significance (p = 1.5 × 10^−7^) driven by several of the same variants. Further examination of results from the single variant tests in *ATG4C* identified a frameshift variant with frequency of 0.002 (1:62834058-TTG-T) - too high to be included in our burden test - that just missed our threshold for testing in the follow-up cohorts (*p* = 0.0003, Beta = 0.55 in the Broad meta-analysis). This variant also showed evidence of association in the meta-analysis of the Sanger cohorts (*p* = 1.3 × 10^−5^), and also exceeded our study-wide significance threshold in the five-way meta-analysis of all cohorts (*p* = 1.55 × 10^−8^). Of further note, an additional *ATG4C* frameshift variant specifically enriched in Finland (1:62819215:C:CT) is associated with IBD (*p* = 6.91 × 10^−8^, Beta = 1.20) in the publicly released FinnGen resource (r5.finngen.fi). All variants in burden and individual testing increase risk, and the inclusion of four truncating variants in these analyses suggests that loss-of-function variants in *ATG4C* strongly increase CD risk.

## Discussion

Here, we demonstrate that large-scale exome sequencing can complement GWAS by pinpointing specific genes both indirectly implicated by GWAS as well as those not yet observed in GWAS. With high sensitivity to directly test individual variants down to 0.01% minor allele frequency, as well as assess burden of ultra-rare mutations, we begin to fill in the low-frequency and rare-variant component of the genetic architecture of Crohn’s disease. This component was not observable by earlier generations of CD GWAS meta-analyses, which have had more limited coverage of low-frequency and rare variation.

Past findings in IBD^5^, and most other complex diseases, suggest that while coding variants are vastly outnumbered by non-coding variation, they are highly enriched for associations to common and rare disease. Furthermore, associated coding variants tend to have stronger effects than their non-coding counterparts, often keeping them lower in frequency via natural selection. While this alone validates the use of exome sequencing for efficiency’s sake, the primary advantage of targeting coding regions for discovery is that coding variants uniquely pinpoint genes, and often pathogenetic mechanisms, in a fashion that is at present far more challenging to achieve routinely for non-coding associations. In the case of several of the new findings (e.g., *RELA, TAGAP*), the coding variation here provides concrete evidence of genes previously indirectly implicated by independent non-coding GWAS associations. These identify the likely gene underlying these associations and build allelic series of natural perturbations at these genes. Moreover, *IL10RA* and *RELA* are known to harbor mutations causing rare, Mendelian, inflammatory GI disorders and this study extends the phenotypic spectrum resulting from their genetic perturbation to more complex forms of CD. From a functional perspective, the novel genes identified in the current study reiterate the central role of innate and adaptive immune cells as well as autophagy in CD pathogenesis. Moreover, the involvement of *PDLIM5* as well as the *CCR7*/*CCL19*/*CCL21* pathway highlights the emerging role of mesenchymal cells in the development and maintenance of intestinal inflammation^18^.

We expect that, in the next year, expanded sequencing efforts underway in ulcerative colitis will come to completion, enabling a more comprehensive survey of low-frequency and rare variation in ulcerative colitis, and IBD in general. Integrated with a much larger GWAS spearheaded in parallel by the IIBDGC, we expect a substantial number of conclusively linked genes and informative allelic series to emerge.

### Box 1.

**Genes newly implicated in Crohn’s disease risk**

#### DOK2

(Docking Protein 2, Downstream of Tyrosine Kinase 2) is a cytoplasmic signaling protein highly expressed in macrophages and T-cells in the terminal ileum. Loss of Dok-2 in mice causes severe DSS-induced colitis with reduced IL-17A and IL-22 expression^19^, and *DOK2* is known to be differentially methylated in colonic tissue of IBD patients^20^. *DOK2* regulates both *TLR2*-induced inflammatory signaling and NK cell development, and *DOK2* loss-of-function is associated with increased IFN-γ production^21,22^. The P274L variant has previously been implicated in atopic eczema where the rare allele was significantly protective for atopic eczema likely by disturbing the RasGAP activation of DOK2 and transcriptomic analyses also suggest that *DOK2* is a central hub interacting with *CD200R1, IL6R*, and *STAT3*^*23*^.

#### TAGAP

(T-Cell Activation RhoGTPase Activating Protein) has a pivotal role in TH17 development and modulates the risk of autoimmunity through influencing thymocyte migration in thymic selection^24,25^. *TAGAP* expression is upregulated in rectal tissue in IBD patients, and *TAGAP* is required for Dectin-induced anti-fungal signaling and proinflammatory cytokine production in myeloid cells^26,27^.

#### PTAFR

(Platelet Activating Factor Receptor), a hypoxia response gene, has an affinity for bacterial phosphorylcholine (ChoP) moieties^28^ and influences development of cigarette-induced Chronic Obstructive Pulmonary Disease (COPD) by inducing neutrophil autophagic death in mice^29^. *PTAFR* regulates colitis-induced pulmonary inflammation through the NLRP3 inflammasome^30^.

#### PDLIM5

(PDZ And LIM Domain 5) is a kidney anion exchanger and scaffolding protein. Genetic variation in this gene is associated with prostate cancer, schizophrenia, diverticular disease, diverticulosis, colorectal cancer, testicular cancer, and self-reported angina^31^. *PDLIM5* has been reported to be a *STAT3* interaction partner involved in actin binding^32^, with *STAT3* previously being identified as an IBD gene^33^. *PDLIM5* is highly expressed in myofibroblast cells, which are important mesenchymal cells of the intestinal lamina propria^18^.

#### SDF2L1

(Stromal Cell Derived Factor 2 Like 1) has been recently identified in the ER stress response in primary intestinal epithelial cells^34^. *SDF2L1* is an ER resident protein that functions within a large protein complex regulating the BiP and Erdj3 chaperone cycle to promote protein folding and secretion^35–37^. Structurally, Sdf2l1 contains an N-terminal signal peptide for entry into the ER lumen and a C-terminal ER retention signal flanking 3 MIR domains that promote complex assembly. The CD risk variant R161H is located in the third MIR domain. In murine models, deletion of *SDF2L1* in the liver resulted in prolonged ER stress and insulin resistance^38^. In the intestine, single cell transcriptional profiling revealed that *SDF2L1* is predominantly expressed in highly secretory cell lineages, including mucin-secreting goblet cells and immunoglobulin-secreting plasma cells^39^. Moreover, *SDF2L1* expression is dynamically regulated and specifically induced during the acute phase of the unfolded protein response (UPR)^34^. Together, these observations suggest a critical role for *SDF2L1* in maintaining ER homeostasis and secretory capacity, which may promote barrier function at the level of mucus integrity and/or neutralization of immunoglobulins and antimicrobial peptides that collectively limit interactions between luminal microbes and the host immune system.

#### CCR7

(chemokine receptor 7) and its ligands *CCL19/CCL21* promote homing of T-cells and dendritic cells to T-cell areas of lymphoid tissues where T-cell priming occurs. *CCR7* also contributes to adaptive immune functions including thymocyte development, secondary lymphoid organogenesis, high affinity antibody responses, regulatory and memory T-cell function, and lymphocyte egress from tissues. *CCR7* expression is upregulated in an inflamed gut in CD^40^, and *CCR7* regulates the intestinal TH1/TH17/Treg balance during Crohn’s-like murine ileitis^41^. Genetic variation in *CCR7* is associated with atopy, asthma, COPD, and IBD in African-Americans^31^. *CCL19* and *CCL21* are highly expressed in a population of stromal cells (designated as S4) that are expanded in IBD inflamed tissues and that continually produce proinflammatory factors preventing the resolution phase of a wound-healing response^18^.

#### IL10RA

(Interleukin 10 receptor A) is a potent regulator of innate and adaptive immune responses, and *IL10RA* genetic variants are associated with Very-Early Onset IBD (VEOIBD) cases; a subset of VEOIBD refractory patients respond to hematopoietic stem cell transplantation^42^. IL10RA (aka *Il10r1)* knockout mice are susceptible to chemical-induced colitis^43^.

#### RELA

(Nuclear Factor NF-Kappa-B P65 Subunit). NFkB is a ubiquitous transcription factor, and its most abundant form is *NFKB1* complexed together with *RELA. RELA* regulates the Th17 pathway in autoimmune disease models^44^, and the FOXO3-NF-κB RelA protein complexes reduce proinflammatory cell signaling and function^45^. *RELA* haploinsufficiency causes autosomal dominant chronic mucotaneous ulceration^46^, and *RELA* is a master transcriptional regulator of epithelial-mesenchymal transition in epithelial cells^47^. Genetic variation in *RELA* has been associated with SLE, type 2 diabetes, psoriasis, obesity, asthma, and atopic dermatitis^31^.

#### ATG4C

(Autophagy-Related 4C Cysteine Peptidase) defective autophagy is established as a mechanism contributing to CD risk. This gene encodes one of four Atg4 isoforms (Atg4A, B, C, and D) that prime pro-LC3 and GABARAP (orthologues of yeast Atg8), essential proteins required for autophagosome biogenesis^48,49^. These Atg4 proteins, including Atg4C, are involved with proteolytic cleavage of Atg8’s C-terminus, thus exposing a specific Atg8 glycine residue necessary for phospholipid covalent binding to Atg8. Atg8 lipidation is necessary for autophagosome formation^50^.

#### HGFAC

Hepatocyte Growth Factor Activator is a serine endopeptidase that converts Hepatocyte Growth Factor (HGF) to its active form in response to thrombin and kallikrein endopeptidases. HGF contributes to neutrophil recruitment. HGF expression is increased in active UC with animal models, suggesting that HGF-MET signaling exacerbates intestinal inflammation^51,52^. Furthermore, HGF promotes colonic epithelial regeneration and mucosal repair^53,54^. *HGFAC* variation is also associated with tuberculosis susceptibility^55^.

## Supplementary Material / Methods

### Broad Institute sequencing pipeline

#### Sample processing

Exome sequencing was performed at the Broad Institute. The sequencing process includes sample prep (Illumina Nextera, IIlumina TruSeq, and Kapa Hyperprep), hybrid capture (Illumina Rapid Capture Enrichment (Nextera) - 37Mb target, and TWIST Custom Capture - 37Mb target), and sequencing (Illumina HiSeq2000, Illumina HiSeq2500, Illumina HiSeq4000, Illumina HiSeqX, Illumina NovaSeq6000 - 76bp and 150bp paired reads). Sequencing was performed at a median depth of 85% targeted bases at >20X. Sequencing reads were mapped by BWA-MEM to the hg38 reference using a ‘functional equivalence’ pipeline. The mapped reads were then marked for duplicates, and base quality scores were recalibrated. They were then converted to CRAMs using Picard and GATK. The CRAMs were then further compressed using ref-blocking to generate gVCFs. These CRAMs and gVCFs were then used as inputs for joint calling. To perform joint calling, the single-sample gVCFs were hierarchically merged (separately for samples using Nextera and Twist exome capture).

#### Quality control

Quality control (QC) analyses were conducted in Hail v0.2.47 (**Supplementary Figure 1**). We first split multiallelic sites and code genotypes with genotype quality (GQ) lower than 20 as missing. Variants not annotated as frameshift, inframe deletion, inframe insertion, stop lost, stop gained, start lost, splice acceptor, splice donor, splice region, missense, or synonymous were removed from the following analysis. We also removed variants that have known quality issues (have a non-empty QUAL column) in the gnomAD dataset. *Sample QC:* poor-quality samples that met the following criteria were identified and removed: 1) samples with an extremely large number of singletons (≥ 500); 2) samples with mean GQ < 40; and 3) samples with missingness rates > 10%. *Variant QC:* low-quality variants that met the following criteria were identified and removed: 1) variants with missingness rate > 5%; 2) variants with mean read depth (DP) < 10; 3) variants that failed the Hardy-Weinberg Equilibrium (HWE) test for controls with *p* < 1 × 10^−4^; and 4) variants with > 10% samples that were heterozygous and with an allelic balance ratio < 0.3 or > 0.7. Variants with different genotypes in WES and WGS in gnomAD were also removed. For Twist exome capture samples, we additionally removed 1) samples that had a significantly high or low inbreeding coefficient (> 0.2 or < -0.2); 2) samples that had a high heterozygosity away from mean (± 5 standard deviations); and 3) related samples, which were removed sequentially by removing the individual with the largest number of related samples (in PLINK, the individual with PI_HAT > 0.2 when using the --genome option) until no related samples remained. For Nextera capture samples, we additionally removed variants showing a significant heterogeneous effect across Ashkenazi Jewish (AJ), Lithuanian (LIT), Finnish (FIN), and non-Finnish European (NFE) samples (see **“Population Assignment”** below).

#### Population assignment

We projected all samples onto principal component (PC) axes generated from the 1000 Genomes Project Phase 3 common variants, and classified their ancestry using a random forest method to the European (CEU, TSI, FIN, GBR, IBS), African (YRI, LWK, GWD, MSL, ESN, ASW, ACB), East Asian (CHB, JPT, CHS, CDX, KHV), South Asian (GIH, PJL, BEB, STU, ITU) and American (MXL, PUR, CLM, PEL) samples. We kept samples that were classified as European with prediction probability greater than 80%. For Nextera samples, we used a second random forest classifier to assign EUR samples to AJ, LIT, FIN, or NFE, and a third random forest classifier to clean the AJ/NFE split.

#### Meta-analysis

We used METAL^56^ with the inverse variance weighted (IVW) fixed-effect model to meta-analyse the SAIGE association statistics from Nextera and Twist samples (**Table 1**). The heterogeneity test was performed using Cochran’s Q with one degree of freedom.

### Sanger Institute sequencing pipeline

#### Sample processing

Genome sequencing was performed at the Sanger Institute using the Illumina HiSeq X platform with a combination of PCR (n=4751, controls only) and PCR-free library preparation protocols. Sequencing was performed at a median depth of 18.6X. Exome sequencing of cases was performed at the Sanger Institute using the Illumina NovaSeq 6000 and the Agilent SureSelect Human All Exon V5 capture set. Controls from the UK Biobank were sequenced separately as a part of the UKBB WES50K release using Illumina NovaSeq and the IDT xGen Exome Research Panel v1.0 capture set (including supplemental probes). 33,704 UKBB participants were selected for use as controls, excluding participants with recorded or self-reported CD, UC, unspecified noninfective gastroenteritis or colitis, any other immune-mediated disorders, or a history of being prescribed any drugs used to treat IBD. Exome and genome datasets were analysed separately but followed a similar analysis protocol.

Reads were mapped to hg38 reference using BWA-MEM. Variant calls were performed using a GATK 4 Best Practices-like pipeline; per-sample intermediate variant calling was followed by joint genotyping across the individual genome and exome cohorts. For the exome cohort, variant calling was limited to Agilent extended target regions. Per-region VCF shards were imported into the Hail software and combined. Multi-allelic sites were split. For the exome cohort, we subsetted the calls to the intersection of Agilent and IDT exome captures, further excluding regions recommended for exclusion by the UKBB due to an error in read mapping that results in no variant calls made.

#### Population assignment

We selected a set of ∼14,000 well-genotyped common variants to identify the genetic ancestry of individual participants through the projection of 1000 Genomes Project cohort-derived principal components. For genomes, due to primarily European genetic ancestry of the controls, we excluded samples outside of four median absolute deviations from the median point of the European ancestry cluster of 1000G. For exomes, we implemented a Random Forest technique that classified samples based on principal components into broad genetic ancestry groups (EUR, AFR, SAS, EAS, admixed), with self-reported ancestry as training labels. For these analyses, we only retained the EUR samples, as the number of cases for other groups was too small for robust association analysis.

#### Quality control

A combination of hard-cutoff filters and per-ancestry/per-batch outlier filters were used to identify low-quality samples. We applied hard-filters for sample depth (> 12x genomes, > 15x exomes), call rate (> 0.95), chimerism < 0.5 (WGS) and FREEMIX < 0.02 (WGS). We excluded genotype calls with an allelic imbalance (for hets, (ab < 0.20) | (ab > 0.80)), low depth (< 2x), and low GQ (< 20). We then performed per-ancestry and per-sequencing protocol (AGILENT vs IDT for WES, PCR vs PCR-free for WES) filtering of samples falling outside 4 MAD from the median per-batch heterozygosity rate, Ti/Tv rate, number of called SNPs and INTELs, and insertion and deletion counts/ratio.

An ancestry-aware relatedness calculation (pc-relate method in Hail^57^) was used to identify related samples. As our association approach (logistic mixed-models) can control for residual relatedness, we only excluded duplicates or MZ twins from within the cohorts and excluded second and third degree relatives when the kinship was across the cohorts (e.g., parent in WGS, child in WES; kinship metric > 0.1 calculated via PC-Relate method using 10 principal components). In addition, we removed samples that were also present in the Broad Institute’s cohorts.

#### Association testing

Association analysis was performed using a logistic mixed-model implemented in the REGENIE software. A set of high-confidence variants (> 1% MAF, 99% call rate, and in Hardy-Weinberg Equilibrium) was used for *t-*fitting. To control for case-control imbalance, Firth correction was applied to *p*-values < 0.05. To control for residual ancestry and sequencing heterogeneity, we calculated 10 principal components on a set of well-genotyped common SNPs, excluding regions with known long-range LD. These were used as covariates for association analyses. Only variants with call-rate above 90% after filtering poor calls were included in the association analysis. For WES, we verified that the > 90% call-rate condition holds true in both AGILENT and IDT samples. Association analysis was performed on QC-passing calls.

### Kiel/Regeneron sequencing pipeline

#### Sample Preparation and Sequencing

The DNA samples were normalized and 100ng of genomic DNA was prepared for exome capture with custom reagents from New England Biolabs, Roche/Kapa, and IDT using a fully-automated approach developed at the Regeneron Genetics Center. Unique, asymmetric 10 base pair barcodes were added to each side of the DNA fragment during library preparation to facilitate multiplexed exome capture and sequencing. Equal amounts of sample were pooled prior to exome capture with a slightly modified version of IDT’s xGen v1 probes; supplemental probes were added to capture regions of the genome well-covered by a previous capture reagent (NimbleGen VCRome) but poorly covered by the standard xGen probes. Captured fragments were bound to streptavidin-conjugated beads and non-specific DNA fragments were removed by a series of stringent washes according to the manufacturer’s recommended protocol (IDT). The captured DNA was PCR amplified and quantified by qRT-PCR (Kapa Biosystems). The multiplexed samples were pooled and then sequenced using 75 bp paired-end reads with two index reads on the Illumina NovaSeq 6000 platform using S2 flow cells.

#### Variant calling and quality control

Sample read mapping and variant calling, aggregation and quality control were performed via the SPB protocol described in Van Hout et al. ^58^. Briefly, for each sample, NovaSeq WES reads are mapped with BWA MEM to the hg38 reference genome. Small variants are identified with WeCall and reported as per-sample gVCFs. These gVCFs are aggregated with GLnexus into a joint-genotyped, multi-sample VCF (pVCF). SNV genotypes with read depth (DP) less than seven and indel genotypes with read depth less than ten are changed to no-call genotypes. After the application of the DP genotype filter, a variant-level allele balance filter is applied, retaining only variants that meet either of the following criteria: (i) at least one homozygous variant carrier or (ii) at least one heterozygous variant carrier with an allele balance (AB) greater than the cutoff.

#### Analysis

We combined the gvcf files with bcftools 1.11 using the “merge” command, then imported the joint vcf into Hail. We then split the multiallelic variants and removed variants with “<NON_REF>“ alternative alleles. We applied the QC steps and assigned populations as in the Broad Institute sequencing pipeline.

#### Cross-cohort meta-analysis

We used the Cochran–Mantel–Haenszel (CMH) test to combine association summary statistics between the Broad Institute, Sanger Institute and Kiel/Regeneron cohorts.

#### Relation to known IBD causal variants

We assigned study-wide significant variants to one of four categories (**Supplementary Table 4**): 1) <u>Known causal candidates</u>: variants in a fine-mapping credible set^5^ with posterior inclusion probability (PIP) > 5%, or reported in the earlier sequencing studies^6,8^. 2) <u>New locus</u>: variants implicating a genetic locus in general onset CD for the first time. 3) <u>Tagging variants</u>: tagging variants with the best PIP in fine-mapping credible sets using conditional analysis (see **“Conditional analysis”** below). 4) <u>New variants in known locus</u>: variants in known GWAS loci but either have MAF < 0.5% (and thus, no LD to evaluate tagging) or remain study-wide significant after conditional analysis using the LD from gnomAD.

#### Conditional analysis

For study-wide significant variants not in a previously reported credible set^5^, we performed a conditional analysis to test whether they are independent from or tagging the known causal variants^5^. We first classified variants as “tagging” if they had *r*^*2*^ > 0.8 with any variants in the reported credible sets^5^. For other variants, we performed a conditional analysis using 1) the *p*-value estimates from previous fine-mapping studies for credible set variants and 2) the LD calculated from gnomAD. We were unable to directly fit a multivariate model or use the LD from study subjects, because exome sequencing does not cover the non-coding putative causal variants, and the ImmunoChip does not have good quality for rare coding variants. The conditional *z* statistic, *z′*_*seq*’_ for a variant with marginal statistic of *z*_*seq*_ from our study, was calculated as follows:

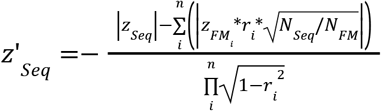

in which 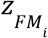 is the *z* statistic of the variant with the best PIP in the credible set *i* from the fine-mapping study, *r*_*i*_ is the LD between the two variants, and *N*_*seq*_ and *N*_*FM*_ are the effective sample sizes for our study and the fine-mapping study respectively. We used the absolute value in this equation because of the challenges to align the alleles across sequencing, the fine-mapping study, and the gnomAD reference panel. Taking the absolute value is a conservative approximation (less likely to declare a variant as novel association) because it assumes that the putative causal variants from fine-mapping have the same direction of effect as the variant being tested when they are in LD. This is very likely to be correct. The effective sample size was calculated as 4/(1/*N*_*case*_ + 1*/N*_*control*_), in which *N*_*case*_ and *N*_*control*_ are the sample sizes for cases and controls respectively. For each variant, we summed the effective sample sizes across all cohorts in which the variant is observed (thus, *N*_*seq*_ can differ from variant to variant). We calculated the conditional *p-*value of *z*′_*seq*_ under the standard Gaussian distribution. A variant was classified as “tagging” if the conditional *p-*value failed to reach study-wide significance at 3 × 10^−7^.

#### Exceptions and notes

*<u>HGFAC</u>*: despite this locus having been reported in an earlier GWAS^2^, the coding variant we identify was missed and is reported in this study for the first time as directly implicating this gene (*r*^*2*^ = 0.35 with the previously reported GWAS SNP, rs2073505). We thus assign this variant as “New variant in known locus”. *<u>RELA</u>*: similarly to *HGFAC*, this locus has been reported in an earlier GWAS^2^, but the coding variant we identify was missed and is reported in this study for the first time as directly implicating this gene (*r*^*2*^ = 0.002 with the previously reported GWAS SNP, rs568617). We thus assign this variant as “New variant in known locus”. *<u>SDF2L1</u>*: this variant has marginal *p*-value = 2 × 10^−7^ and conditional *p* = 3.4 × 10^−4^. The *r*^*2*^ between this variant and the non-coding variant with the best PIP from fine-mapping is 0.045. We manually assigned this variant to “New variants in known locus,” as this is a missense variant. *<u>SLC39A8</u>*: the *SLC39A8* A391T variant was not reported in the fine-mapping paper, as its genetic region was not included in the ImmunoChip design. Because this variant has been published in several papers as an IBD variant with genetic and functional evidence^59–61^, we assign this variant as “Known causal candidate”. *<u>TYK2</u>*: the *TYK2* A928V was not reported in the fine-mapping paper^5^, likely due to a lack of power. Because this variant has been known to be a causal variant for several autoimmune disorders^62^ and in another IBD study^63^, we assign this variant as “Known causal candidate”. *<u>NOD2</u>*: a) Previous studies^5–7^ have shown evidence that the *NOD2* S431L variant tags the *NOD2* V793M variant, with the latter more likely to be the CD causal variant. In this study, however, S431L reached study-wide significance, but V793M failed to meet the significance cutoff. We therefore retained S431L in **Figure 1** for the purpose of keeping this association signal. b) Due to the complexity of the *NOD2* locus, we conducted a haplotype analysis using the Twist subjects and additionally classified signed variants that share the same haplotype with known IBD variants as “tagging”. We found that for the *NOD2* S47L variant, 18 out of 19 copies of the T allele are on the same haplotype as the fs1007insC variant. We therefore classify S47L as “tagging”. c) The NOD2 A755V variant is in LD with rs184788345, the best PIP variant from fine-mapping (*r*^*2*^ = 0.85). The marginal *p-*value for A755V is one order of magnitude less significant than rs184788345. Considering A755V is a missense variant while none of the variants in the credible set defined by rs184788345 are coding, we assign A755V as a likely ROse”Known causal candidate”.

## Data Availability

Sequence data is publicly available in dbGaP Study Accession: phs001642.v1.p1 - Center for Common Disease Genomics [CCDG] - Autoimmune: Inflammatory Bowel Disease (IBD) Exomes and Genomes (https://www.ncbi.nlm.nih.gov/projects/gap/cgi-bin/study.cgi?study_id=phs001642.v1.p1).
The summary statistics of Nextera and TWIST meta-analysis have been deposited on GitHub (https://github.com/yorkklause/Crohn-s-Disease-WES-meta).

## Data availability

Sequence data has been made publicly available in dbGaP Study Accession: phs001642.v1.p1 - Center for Common Disease Genomics [CCDG] - Autoimmune: Inflammatory Bowel Disease (IBD) Exomes and Genomes (https://www.ncbi.nlm.nih.gov/projects/gap/cgi-bin/study.cgi?study_id=phs001642.v1.p1).

The summary statistics of Nextera and TWIST meta-analysis have been deposited on GitHub (https://github.com/yorkklause/Crohn-s-Disease-WES-meta).

## Acknowledgements

We thank all the principal investigators, local staff from individual cohorts, and all of the patients who kindly donated samples used in the study for making possible this global collaboration and resource to advance IBD genetics research. This research was funded in whole, or in part, by the US National Institutes of Health Grants U54HG003067 and 5UM1HG008895, the Wellcome Trust Grant 206194, and The Leona M. & Harry B. Helmsley Charitable Trust 2015PG-IBD001. We thank the Broad Institute Genomics Platform for genomic data generation efforts and the Stanley Center for Psychiatric Research at the Broad Institute for supporting control sample aggregation. M.A.R. is in part supported by the NHGRI of the NIH under award R01HG010140 (M.A.R.) and an NIH Center for Multi- and Trans-ethnic Mapping of Mendelian and Complex Diseases grant (5U01HG009080). H.H. acknowledges support from NIDDK K01DK114379 and the Stanley Center for Psychiatric Research. H.H.U. and A.S. are supported by the NIHR Oxford Biomedical Research Centre and by The Leona M. and Harry B. Helmsley Charitable Trust.

Individual studies contributing to this meta-analysis acknowledge support from NIH grants DK062431, DK062432, DK087694, K23DK117054, R01DK111843, P01DK094779, R01HG010140, 5U01HG009080, and DK062420, National Institute of Diabetes and Digestive and Kidney Disease (NIDDK) Grants P01DK046763, U01DK062413, and R01DK104844. We acknowledge the Crohn’s & Colitis Foundation’s IBD Plexus Program - the results published here are in part based on data obtained from the IBD Plexus program of the Crohn’s & Colitis Foundation. We acknowledge the contribution of the Oxford IBD cohort study and the Oxford GI biobank, which are supported by the NIHR Oxford Biomedical Research Centre. We acknowledge individual study support from FNRS (Fond National de la Recherche Scientifique, Belgium), the NIHR Newcastle Biomedical Research Centre, the Charles Wolfson Charitable Trust, the Canadian Institute for Health Research Grants SPOR-RN279389–358033, the Academy of Finland Centre of Excellence in Complex Disease Genetics (312074, 336824), the German Excellence Initiative EXC 2167 and IMI Programm of the EU (“3TR”), and the European Regional Development Fund 01.2.2-LMT-K-718-04-0003. H.S.W. receives philanthropic support from Martin Schlaff, James Brooks, and the B. Hasso Family Foundation. The German IBD Consortium would like to acknowledge the CCIM that receives infrastructure support from the DFG Cluster of Excellence “Precision Medicine in Chronic Inflammation” (PMI). The Cedars-Sinai IBD study was supported by the Cedars-Sinai MIRIAD IBD Biobank. The MIRIAD IBD Biobank receives funding from the Widjaja Foundation Inflammatory Bowel and Immunobiology Research Institute, National Institute of Diabetes and Digestive and Kidney Disease Grants P01DK046763 and U01DK062413, and The Leona M. and Harry B. Helmsley Charitable Trust. The FINRISK controls were part of the FINRISK studies supported by THL (formerly KTL: National Public Health Institute) through budgetary funds from the government, with additional funding from institutions such as the Academy of Finland, the European Union, ministries and national and international foundations and societies to support specific research purposes. The UK/IRL controls were collected and sequenced with support from the Neuroscience Research Charitable Trust, the Central London NHS (National Health Service) Blood Transfusion Service and the National Institute of Health Research (NIHR) funded Mental Health Research Network. The Epi25 Collaborative was supported by the NHGRI Centers for Common Disease Genomics Genomics (CCDG) grant (UM1HG008895) and the Stanley Center for Psychiatric Research. The NIMH Controls were supported by a cooperative agreement grant funded by the National Institute of Mental Health (U01MH100233, U01MH100209, U01MH100229, U01MH100239).

This research has been conducted using the UK Biobank Resource and controls made publically available by dbGaP (phs001000.v1.p1, phs000806.v1.p1 - Myocardial Infarction Genetics Consortium (MIGen), phs000401.v1.p1 - NHLBI GO-ESP Project, phs000298.v4.p3 - Autism Sequencing Consortium (ASC), phs000572.v8.p4 - Alzheimer’s Disease Sequencing Project (ADSP), phs001489.v1.p1 - Epi25 Consortium, phs001095.v1.p1 - T2D-GENES) as well as additional controls from the 1000 Genomes Project, the Epi25 Collaborative, UK-Ireland Collaborators (A. McQuillin, D. Blackwood, A. McIntosh), and collaborators A. Pulver, H. Ostrer, D. Chung, M. Hiltunen, and A. Palotie (H2000 and SUPER cohorts) (**Supplementary Table 1**).

Participants in the INTERVAL randomised controlled trial were recruited with the active collaboration of NHS Blood and Transplant England (www.nhsbt.nhs.uk [nhsbt.nhs.uk]), which has supported field work and other elements of the trial. We thank Klaudia Walter and Kousik Kundu for help with INTERVAL WGS quality control. DNA extraction and genotyping were co-funded by the National Institute for Health Research (NIHR), the NIHR BioResource (http://bioresource.nihr.ac.uk [bioresource.nihr.ac.uk]) and the NIHR Cambridge Biomedical Research Centre (BRC-1215-20014)[*]. Sequencing was supported by the Wellcome Trust grant number 206194. The academic coordinating centre for INTERVAL was supported by core funding from the: NIHR Blood and Transplant Research Unit in Donor Health and Genomics (NIHR BTRU-2014-10024), UK Medical Research Council (MR/L003120/1), British Heart Foundation (SP/09/002; RG/13/13/30194; RG/18/13/33946) and NIHR Cambridge BRC (BRC-1215-20014) [*]. A complete list of the investigators and contributors to the INTERVAL trial is provided in reference [**]. The academic coordinating centre would like to thank blood donor centre staff and blood donors for participating in the INTERVAL trial.

This work was supported by Health Data Research UK, which is funded by the UK Medical Research Council, Engineering and Physical Sciences Research Council, Economic and Social Research Council, Department of Health and Social Care (England), Chief Scientist Office of the Scottish Government Health and Social Care Directorates, Health and Social Care Research and Development Division (Welsh Government), Public Health Agency (Northern Ireland), British Heart Foundation and Wellcome.

*The views expressed are those of the author(s) and not necessarily those of the NIHR or the Department of Health and Social Care.

**Di Angelantonio E, Thompson SG, Kaptoge SK, Moore C, Walker M, Armitage J, Ouwehand WH, Roberts DJ, Danesh J, INTERVAL Trial Group. Efficiency and safety of varying the frequency of whole blood donation (INTERVAL): a randomised trial of 45 000 donors. Lancet. 2017 Nov 25;390(10110):2360-2371.

## SUPPLEMENTARY MATERIALS

**Supplementary Table 1. Sample Cohorts**.

**Supplementary Table 2. Heterogeneity test between samples sequenced using Nextera and Twist capture kits**.

**Supplementary Table 3. 116 variants advanced for replication with the Sanger and Regeneron subjects**.

**Supplementary Table 4. 43 exome-wide significant variants assigned to known/novel categories**.

**Supplementary Table 5. Variants significantly associated with CD in *NOD2***.

**Supplementary Table 6. SAIGE-GENE results**.

**Supplementary Table 7. Individual rare variants driving the *ATG4C* association in four cohorts**.

**Supplementary Figure 1.**
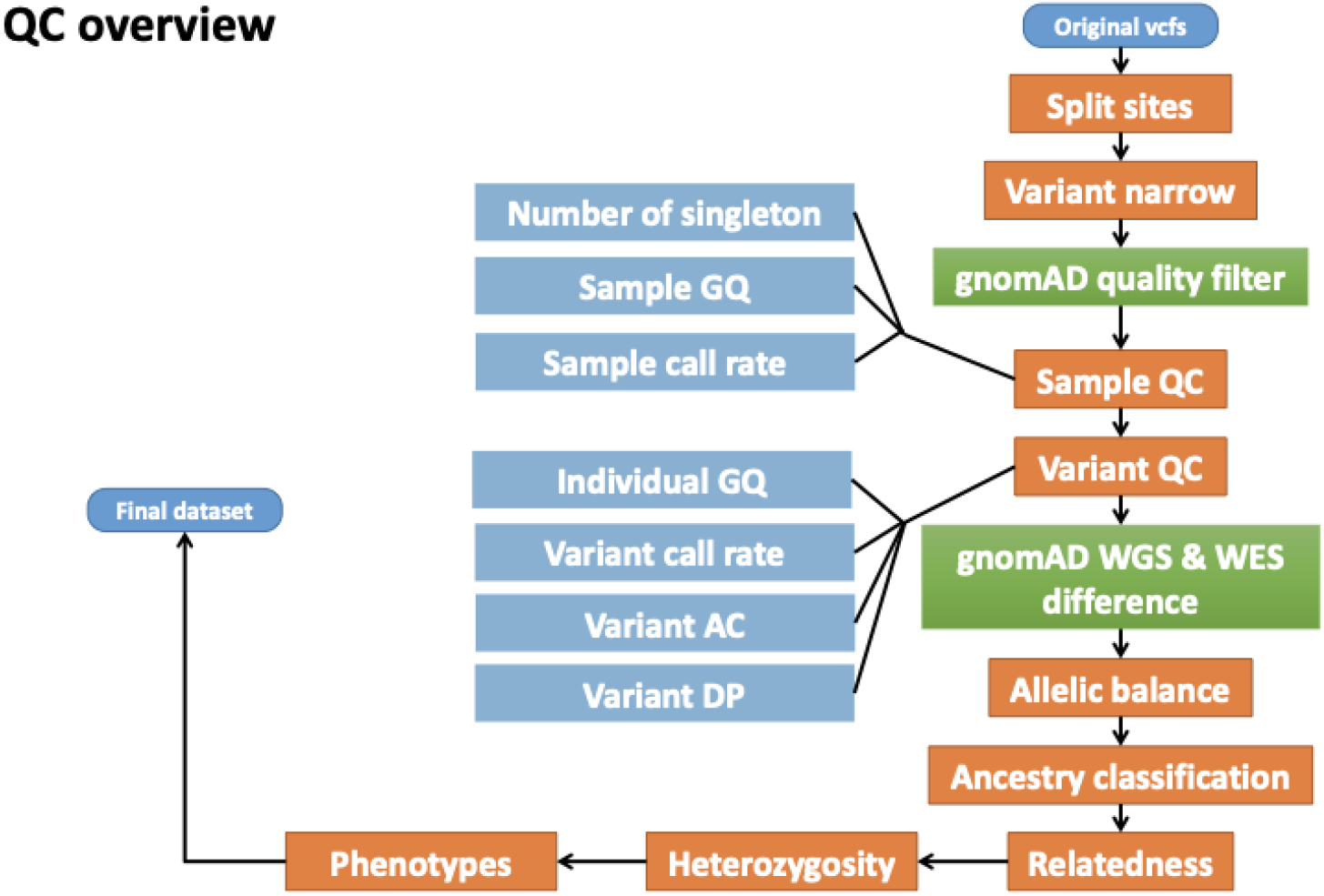
Quality control procedures applied in the Broad sequencing pipeline.

**Supplementary Figure 2.**
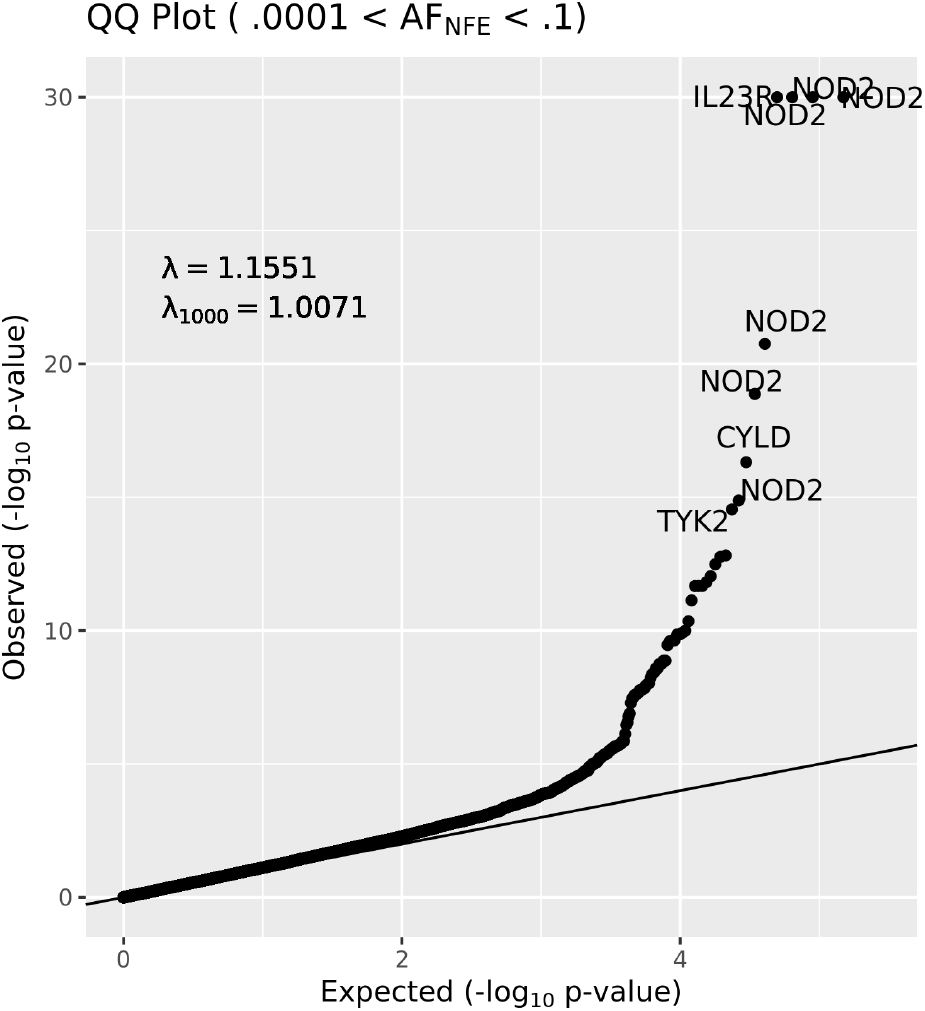
QQ plot of all variants with 0.0001 < MAF < 0.10 passing QC & heterogeneity test from combined Nextera+TWIST. N.b., figure is capped at -log_10_ *p* = 30; the top four variants (the three common *NOD2* and one *IL23R*) have -log_10_ *p* > 100.

## References

1. Jostins, L. et al. Host-microbe interactions have shaped the genetic architecture of inflammatory bowel disease. Nature 491, 119–124 (2012).

2. Liu, J. Z. et al. Association analyses identify 38 susceptibility loci for inflammatory bowel disease and highlight shared genetic risk across populations. Nat. Genet. 47, 979–986 (2015).

3. Luo, Y. et al. Exploring the genetic architecture of inflammatory bowel disease by whole-genome sequencing identifies association at ADCY7. Nat. Genet. 49, 186–192 (2017).

4. de Lange, K. M. et al. Genome-wide association study implicates immune activation of multiple integrin genes in inflammatory bowel disease. Nat. Genet. 49, 256–261 (2017).

5. Huang, H. et al. Fine-mapping inflammatory bowel disease loci to single-variant resolution. Nature 547, 173–178 (2017).

6. Rivas, M. A. et al. Deep resequencing of GWAS loci identifies independent rare variants associated with inflammatory bowel disease. Nat. Genet. 43, 1066–1073 (2011).

7. Rivas, M. A. et al. A protein-truncating R179X variant in RNF186 confers protection against ulcerative colitis. Nature Communications vol. 7 (2016).

8. Rivas, M. A. et al. Insights into the genetic epidemiology of Crohn’s and rare diseases in the Ashkenazi Jewish population. PLoS Genet. 14, e1007329 (2018).

9. Beaudoin, M. et al. Deep resequencing of GWAS loci identifies rare variants in CARD9, IL23R and RNF186 that are associated with ulcerative colitis. PLoS Genet. 9, e1003723 (2013).

10. Cao, Z. et al. Ubiquitin Ligase TRIM62 Regulates CARD9-Mediated Anti-fungal Immunity and Intestinal Inflammation. Immunity 43, 715–726 (2015).

11. Leshchiner, E. S. et al. Small-molecule inhibitors directly target CARD9 and mimic its protective variant in inflammatory bowel disease. Proc. Natl. Acad. Sci. U. S. A. 114, 11392–11397 (2017).

12. Sivanesan, D. et al. IL23R (Interleukin 23 Receptor) Variants Protective against Inflammatory Bowel Diseases (IBD) Display Loss of Function due to Impaired Protein Stability and Intracellular Trafficking. J. Biol. Chem. 291, 8673–8685 (2016).

13. Mohanan, V. et al. C1orf106 is a colitis risk gene that regulates stability of epithelial adherens junctions. Science 359, 1161–1166 (2018).

14. Zhou, W. et al. Efficiently controlling for case-control imbalance and sample relatedness in large-scale genetic association studies. Nat. Genet. 50, 1335–1341 (2018).

15. Festen, E. A. M. et al. A meta-analysis of genome-wide association scans identifies IL18RAP, PTPN2, TAGAP, and PUS10 as shared risk loci for Crohn’s disease and celiac disease. PLoS Genet. 7, e1001283 (2011).

16. Chamaillard, M. et al. Gene–environment interaction modulated by allelic heterogeneity in inflammatory diseases. Proc. Natl. Acad. Sci. U. S. A. 100, 3455–3460 (2003).

17. Zhou, W. et al. Scalable generalized linear mixed model for region-based association tests in large biobanks and cohorts. doi:10.1101/583278.

18. Kinchen, J. et al. Structural Remodeling of the Human Colonic Mesenchyme in Inflammatory Bowel Disease. Cell 175, 372–386.e17 (2018).

19. Waseda, M., Arimura, S., Shimura, E., Nakae, S. & Yamanashi, Y. Loss of Dok-1 and Dok-2 in mice causes severe experimental colitis accompanied by reduced expression of IL-17A and IL-22. Biochem. Biophys. Res. Commun. 478, 135–142 (2016).

20. Cooke, J. et al. Mucosal genome-wide methylation changes in inflammatory bowel disease. Inflamm. Bowel Dis. 18, 2128–2137 (2012).

21. Rhodes, J. Erythrocyte rosettes provide an analogue for Schiff base formation in specific T cell activation. J. Immunol. 145, 463–469 (1990).

22. Celis-Gutierrez, J. et al. Dok1 and Dok2 proteins regulate natural killer cell development and function. EMBO J. 33, 1928–1940 (2014).

23. Mucha, S. et al. Protein-coding variants contribute to the risk of atopic dermatitis and skin-specific gene expression. J. Allergy Clin. Immunol. 145, 1208–1218 (2020).

24. Tamehiro, N. et al. T-cell activation RhoGTPase-activating protein plays an important role in TH17-cell differentiation. Immunol. Cell Biol. 95, 729–735 (2017).

25. Duke-Cohan, J. S. et al. Regulation of thymocyte trafficking by Tagap, a GAP domain protein linked to human autoimmunity. Sci. Signal. 11, (2018).

26. Medrano, L. M. et al. Expression patterns common and unique to ulcerative colitis and celiac disease. Ann. Hum. Genet. 83, 86–94 (2019).

27. Chen, J. et al. TAGAP instructs Th17 differentiation by bridging Dectin activation to EPHB2 signaling in innate antifungal response. Nat. Commun. 11, 1913 (2020).

28. Clark, S. E. & Weiser, J. N. Microbial modulation of host immunity with the small molecule phosphorylcholine. Infect. Immun. 81, 392–401 (2013).

29. Lv, X.-X. et al. Cigarette smoke promotes COPD by activating platelet-activating factor receptor and inducing neutrophil autophagic death in mice. Oncotarget 8, 74720–74735 (2017).

30. Liu, G. et al. Platelet activating factor receptor regulates colitis-induced pulmonary inflammation through the NLRP3 inflammasome. Mucosal Immunol. 12, 862–873 (2019).

31. Ochoa, D. et al. Open Targets Platform: supporting systematic drug–target identification and prioritisation. Nucleic Acids Res. 49, D1302–D1310 (2020).

32. Blumert, C. et al. Analysis of the STAT3 interactome using in-situ biotinylation and SILAC. J. Proteomics 94, 370–386 (2013).

33. Barrett, J. C. et al. Genome-wide association defines more than 30 distinct susceptibility loci for Crohn’s disease. Nat. Genet. 40, 955–962 (2008).

34. You, K. et al. QRICH1 dictates the outcome of ER stress through transcriptional control of proteostasis. Science 371, (2021).

35. Fujimori, T. et al. Endoplasmic reticulum proteins SDF2 and SDF2L1 act as components of the BiP chaperone cycle to prevent protein aggregation. Genes Cells 22, 684–698 (2017).

36. Meunier, L., Usherwood, Y.-K., Chung, K. T. & Hendershot, L. M. A subset of chaperones and folding enzymes form multiprotein complexes in endoplasmic reticulum to bind nascent proteins. Mol. Biol. Cell 13, 4456–4469 (2002).

37. Hanafusa, K., Wada, I. & Hosokawa, N. SDF2-like protein 1 (SDF2L1) regulates the endoplasmic reticulum localization and chaperone activity of ERdj3 protein. J. Biol. Chem. 294, 19335–19348 (2019).

38. Sasako, T. et al. Hepatic Sdf2l1 controls feeding-induced ER stress and regulates metabolism. Nat. Commun. 10, 947 (2019).

39. Smillie, C. S. et al. Intra- and Inter-cellular Rewiring of the Human Colon during Ulcerative Colitis. Cell 178, 714–730.e22 (2019).

40. Autschbach, F., Funke, B., Katzenmeier, M. & Gassler, N. Expression of chemokine receptors in normal and inflamed human intestine, tonsil, and liver--an immunohistochemical analysis with new monoclonal antibodies from the 8th international workshop and conference on human leucocyte differentiation antigens. Cell. Immunol. 236, 110–114 (2005).

41. McNamee, E. N. et al. Chemokine receptor CCR7 regulates the intestinal TH1/TH17/Treg balance during Crohn’s-like murine ileitis. J. Leukoc. Biol. 97, 1011–1022 (2015).

42. Murugan, D. et al. Very early onset inflammatory bowel disease associated with aberrant trafficking of IL-10R1 and cure by T cell replete haploidentical bone marrow transplantation. J. Clin. Immunol. 34, 331–339 (2014).

43. Pils, M. C. et al. Monocytes/macrophages and/or neutrophils are the target of IL-10 in the LPS endotoxemia model. Eur. J. Immunol. 40, 443–448 (2010).

44. Qu, X. et al. TLR4-RelA-miR-30a signal pathway regulates Th17 differentiation during experimental autoimmune encephalomyelitis development. J. Neuroinflammation 16, 183 (2019).

45. Thompson, M. G. et al. FOXO3-NF-κB RelA Protein Complexes Reduce Proinflammatory Cell Signaling and Function. J. Immunol. 195, 5637–5647 (2015).

46. Badran, Y. R. et al. Human RELA haploinsufficiency results in autosomal-dominant chronic mucocutaneous ulceration. J. Exp. Med. 214, 1937–1947 (2017).

47. Tian, B. et al. The NFκB subunit RELA is a master transcriptional regulator of the committed epithelial-mesenchymal transition in airway epithelial cells. J. Biol. Chem. 293, 16528–16545 (2018).

48. Rioux, J. D. et al. Genome-wide association study identifies new susceptibility loci for Crohn disease and implicates autophagy in disease pathogenesis. Nat. Genet. 39, 596–604 (2007).

49. McCarroll, S. A. et al. Deletion polymorphism upstream of IRGM associated with altered IRGM expression and Crohn’s disease. Nat. Genet. 40, 1107–1112 (2008).

50. Agrotis, A., Pengo, N., Burden, J. J. & Ketteler, R. Redundancy of human ATG4 protease isoforms in autophagy and LC3/GABARAP processing revealed in cells. Autophagy 15, 976–997 (2019).

51. Finisguerra, V. et al. MET is required for the recruitment of anti-tumoural neutrophils. Nature 522, 349–353 (2015).

52. Stakenborg, M. et al. Neutrophilic HGF-MET signaling exacerbates intestinal inflammation. J. Crohns. Colitis (2020) doi:10.1093/ecco-jcc/jjaa121.

53. Kanayama, M. et al. Hepatocyte growth factor promotes colonic epithelial regeneration via Akt signaling. Am. J. Physiol. Gastrointest. Liver Physiol. 293, G230–9 (2007).

54. Tahara, Y. et al. Hepatocyte growth factor facilitates colonic mucosal repair in experimental ulcerative colitis in rats. J. Pharmacol. Exp. Ther. 307, 146–151 (2003).

55. Qi, H. et al. Discovery of susceptibility loci associated with tuberculosis in Han Chinese. Hum. Mol. Genet. 26, 4752–4763 (2017).

56. Willer, C. J., Li, Y. & Abecasis, G. R. METAL: fast and efficient meta-analysis of genomewide association scans. Bioinformatics 26, 2190–2191 (2010).

57. Conomos, M. P., Reiner, A. P., Weir, B. S. & Thornton, T. A. Model-free Estimation of Recent Genetic Relatedness. Am. J. Hum. Genet. 98, 127–148 (2016).

58. Van Hout, C. V. et al. Exome sequencing and characterization of 49,960 individuals in the UK Biobank. Nature 586, 749–756 (2020).

59. Nakata, T. et al. A missense variant in SLC39A8 confers risk for Crohn’s disease by disrupting manganese homeostasis and intestinal barrier integrity. Proc. Natl. Acad. Sci. U. S. A. 117, 28930–28938 (2020).

60. Li, D. et al. A Pleiotropic Missense Variant in SLC39A8 Is Associated With Crohn’s Disease and Human Gut Microbiome Composition. Gastroenterology vol. 151 724–732 (2016).

61. Sunuwar, L. et al. Pleiotropic ZIP8 A391T implicates abnormal manganese homeostasis in complex human disease. JCI Insight 5, (2020).

62. Ellinghaus, D. et al. Analysis of five chronic inflammatory diseases identifies 27 new associations and highlights disease-specific patterns at shared loci. Nat. Genet. 48, 510–518 (2016).

63. Diogo, D. et al. TYK2 protein-coding variants protect against rheumatoid arthritis and autoimmunity, with no evidence of major pleiotropic effects on non-autoimmune complex traits. PLoS One 10, e0122271 (2015).

