## Supplementary figures and images for "Sequencing of over 100,000 individuals identifies multiple genes and rare variants associated with Crohns disease susceptibility"

### SF1

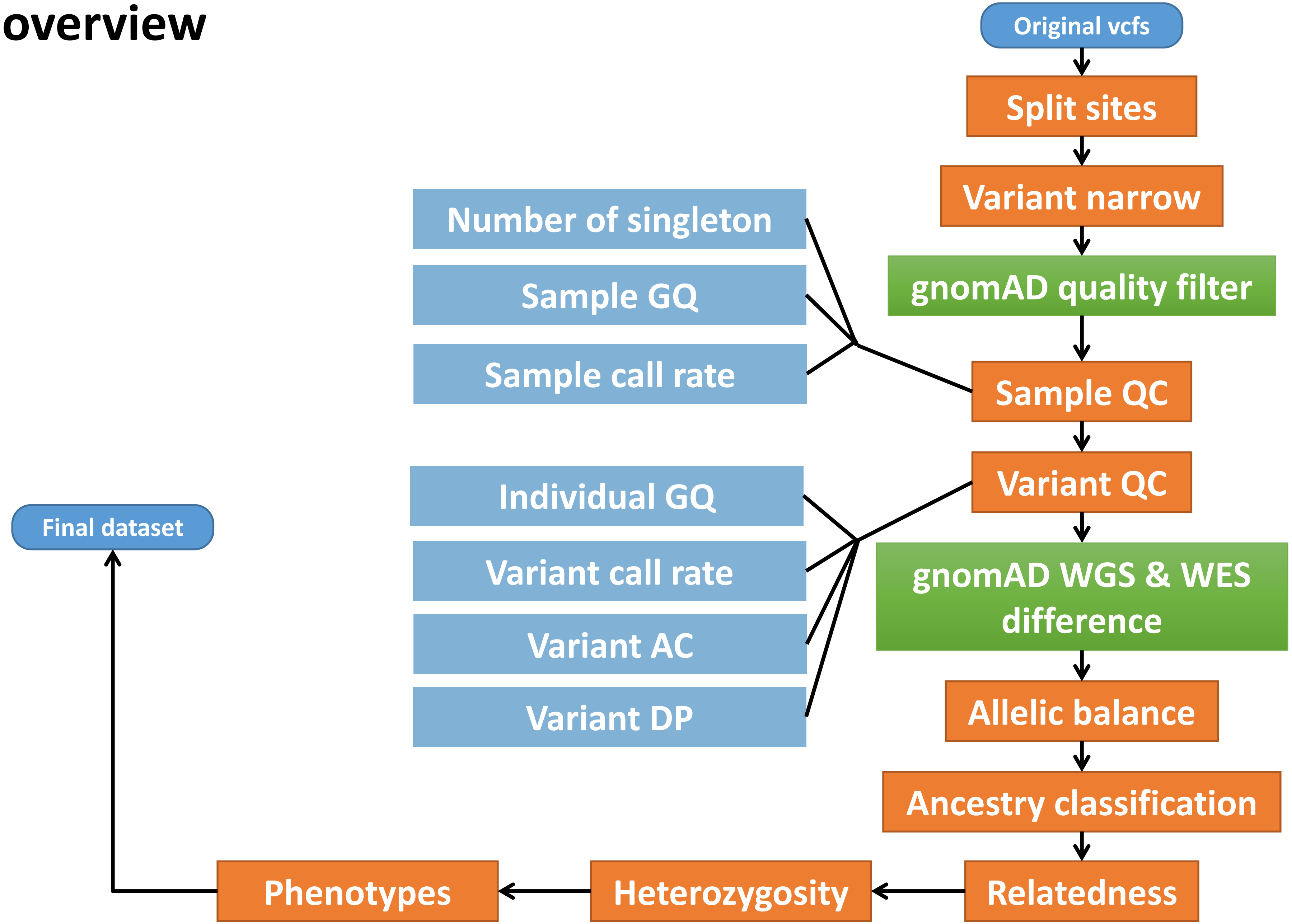

### SF2.tiff

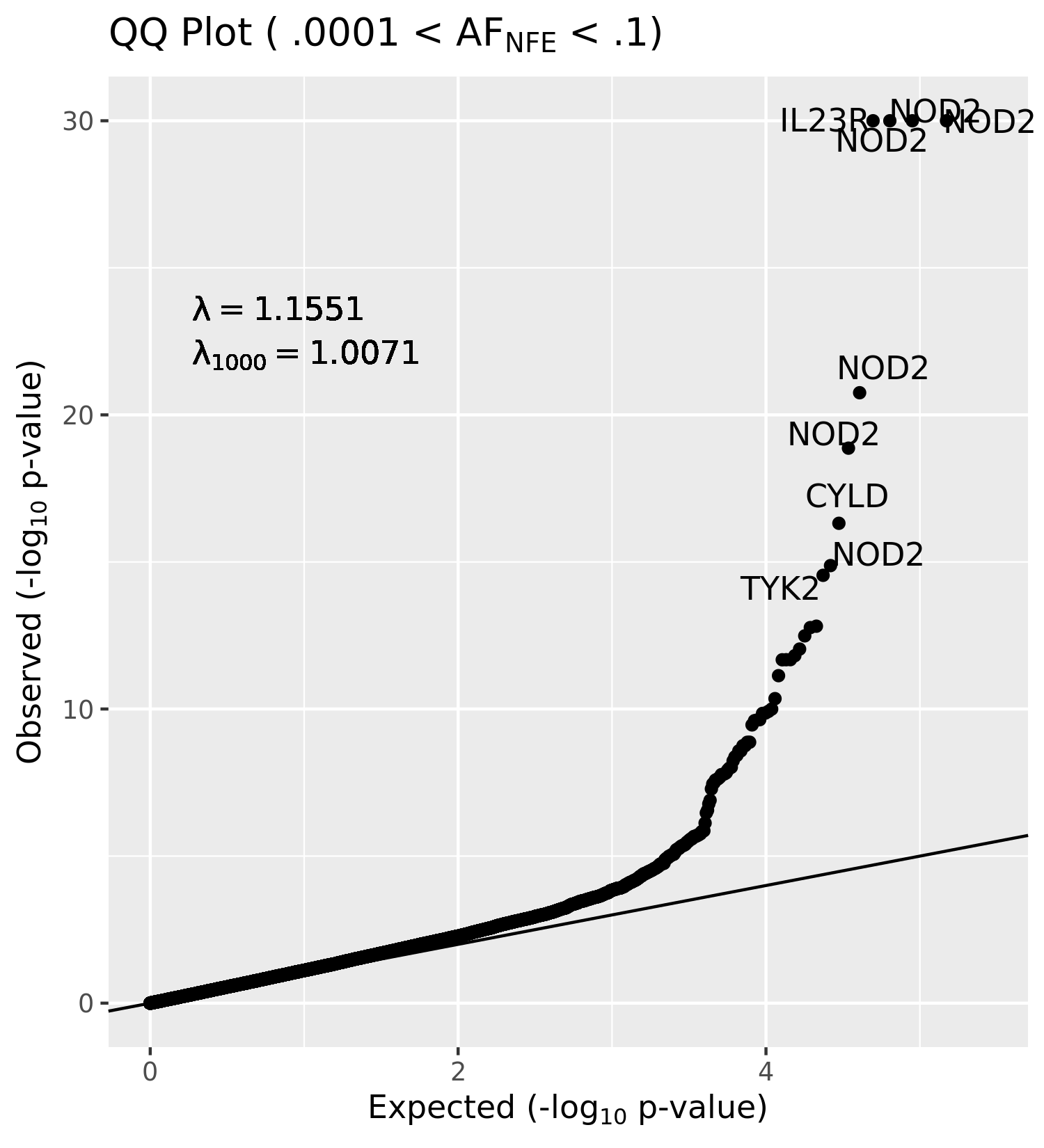
